# Association between grade of congenital hydronephrosis and time to natural resolution

**DOI:** 10.1101/2025.11.26.25341080

**Authors:** V Bleau, A Tsampalieros, N Barrowman, J Seymour, I Terekhov, J Feber, RL Myette

**Author notes:** **Corresponding Author:** Robert L. Myette, Children’s Hospital of Eastern Ontario, University of Ottawa, 401 Smyth Road, Ottawa, Ontario, Canada.

## Abstract

**Introduction:** Standard of care for pregnancy includes a second trimester ultrasound which may lead to incidental findings of fetal uropathies. The most common fetal uropathy is congenital hydronephrosis (CH). There is limited information regarding the natural history of CH. The objective of this study was to describe factors associated with time to natural resolution of CH.

**Methodology:** This is a retrospective cohort study of infants with CH using data from a single hospital’s electronic medical record. Between January 2017 and December 2022, infants with CH were identified using ICD-10 codes and were included in the cohort if they had a second ultrasound prior to 2022. Severity of CH was classified using Society for Fetal Urology (SFU) grades. Cox proportional hazards analysis was used to model time to resolution adjusting for age, sex, and initial SFU grade.

**Results:** Of 209 infants with CH, 126 were included in the cohort, with males predominating (78%). A total of 168 kidney units were included with eighty-four infants (67%) having only 1 kidney unit affected. In multivariable Cox proportional hazards regression modelling, after adjusting for sex and age at initial ultrasound, initial SFU grade was associated with time to natural resolution (compared to grade 1, adjusted hazard ratios for grades 2, 3 and 4 were: 0.59 [95% CI 0.34-1.01], 0.23 [95% CI 0.12,0.47] and 0.04 [95% CI 0.01,0.14]; p<0.001).

**Conclusion:** On average, kidney units with not only SFU 4, but also SFU 3, resolved more slowly than those with low-grade hydronephrosis. These findings will assist in counseling parents of children with CH.

## Introduction

Standard of care for pregnant women includes an ultrasound during the second trimester to document fetal anatomy, which can lead to the antenatal diagnosis of fetal uropathies [1]. The most common fetal kidney anomaly detected on prenatal ultrasound screening is congenital hydronephrosis (CH) with an estimated incidence of ∼5% [2, 3]. The differential diagnosis includes ureteropelvic junction obstruction, ureterovesicular junction obstruction, megacystis megaureter, posterior urethral valves or vesicoureteral reflux [4]. However, there is limited information regarding the natural history of CH and the time to resolution of these lesions [4]. The main challenge is to differentiate CH that will resolve spontaneously postnatally from a pathological obstruction that would require intervention [3, 5]. Early recognition of pathological obstruction is crucial as it can result in significant kidney impairment with long-term consequences if left untreated [2, 3, 5]. The goal is to correctly identify patients who will need early intervention while kidney impairment is still reversible or minimal and spare others from unnecessary invasive procedures [3, 5].

There remain conflicting data in the literature as to which factors are associated with resolution, thus creating uncertainty in the management of infants with CH [6]. Some studies reported a negative correlation between the SFU grade and the likelihood of resolution [2, 4]. Other studies concluded that nonoperative management with close observation for high-grade hydronephrosis was found to be safe with resolution of some of these lesions [1, 6, 7, 8]. Given these discrepant findings, a clear knowledge gap, and the potential for morbidity with surgical intervention, the goal objective of this study was to describe factors associated with time to natural resolution.

## Methodology

### Study Cohort

This study is a retrospective cohort study of infants with CH. This study was approved by The Children’s Hospital of Eastern Ontario (CHEO) research ethics board (REB# 22/38X). Infants with CH were identified from outpatient visits, admissions, and ED visits in the EPIC electronic medical record. Using the Data Warehouse at CHEO, infants with CH were identified using the appropriate ICD code ‘congenital hydronephrosis (ICD-10-CA: Q62.0)’. Manual chart review was performed to ensure that patients with obstructive uropathies not consistent with CH were identified and excluded. Patients were included in the study if they had at least 2 ultrasounds between January 2017 and December 2022 with their first ultrasound within the first month of life. Infants were excluded if their second ultrasound was performed > 14 months after their first kidney ultrasound or if another pathology was identified such as kidney dysplasia or duplex collecting systems. Infants found to have posterior urethral valves on voiding cystourethrogram (VCUG) were also excluded as this pathology would necessitate immediate surgical correction.

## Outcome measurement and covariates of interest

### Hydronephrosis Classification

Severity of CH was classified using the Society of Fetal Urology (SFU) grades. Kidney units were included if the SFU grade was ≥1 at initial measurement. Ultrasound findings between two grades were rounded to the higher SFU grade (for example, an ultrasound finding of 1-2 would be coded as 2). Kidney units were classified as either low-grade hydronephrosis (grade 1 and 2) or high-grade hydronephrosis (grade 3 and 4). The primary outcome was time to resolution of CH defined as SFU grade 0. Individuals who did not resolve during the study period were treated as censored observations. Individuals for whom surgical intervention (such as pyeloplasty, ureteral reimplantation and nephrectomy) was performed were also censored.

### Statistical Analysis

Descriptive statistics were used to summarize results at the patient and kidney unit level. All statistical analyses were performed using the R statistical programming language (Version 4.2.1, [8]). Patient and kidney unit characteristics are reported using frequencies for categorical variables. For continuous variables with skewed distributions, median and interquartile range (IQR) are reported while for normally distributed variables, mean and standard deviation (SD) are reported. Kidney units were censored at time of surgery (if required) or if lost to follow up. A time-to-event analysis was used to model time to resolution with censoring of individuals no longer being followed as well as individuals who underwent surgical intervention. Kaplan-Meier survival curves were used to illustrate the cumulative probability of CH not resolving. Log rank testing was used to compare time to resolution by initial SFU grade (1-2 vs 3-4). In order to estimate the hazard ratio associated with SFU grades 2,3,4 compared to 1 at initial ultrasound, adjusting for age at the time of the first ultrasound and sex, Cox proportional hazard analysis was performed at the kidney unit level. A sensitivity analysis was performed excluding kidney units that underwent surgical intervention. We also conducted a sensitivity analysis in which SFU grades reported as a range (example SFU grade 1-2) were rounded down to the lower SFU grade.

## Results

### Patient Cohort

There were 209 participants screened for CH (Figure 1) with 126 ultimately being included in the study (168 kidney units; Table 1). The median age of infants at first ultrasound was 12 days old (IQR 8,23). Of the children enrolled, 98/126 (78%) were male and 28/126 (22%) were female. The majority of infants had unilateral hydronephrosis with 84/126 (67%) of participants having one kidney (one kidney unit) affected and 42/126 (33%) of participants having both kidneys affected (2 kidney units). There were 105/168 (62%) left kidney units and 63/168 (38%) right kidney units affected by CH. There were 93/168 (55%) kidney units classified as low-grade hydronephrosis (SFU grade 1-2) and 75/168 (45%) kidney units classified as high-grade hydronephrosis (SFU grade 3-4). There were 9 patients (12 kidney units) who underwent surgery. Surgical intervention included either pyeloplasty or ureteral reimplantation. There was one patient who underwent nephrectomy and had a history of ureterocele. The majority of kidney units that underwent surgical intervention were classified as high-grade (11/12).

**Figure 1.**
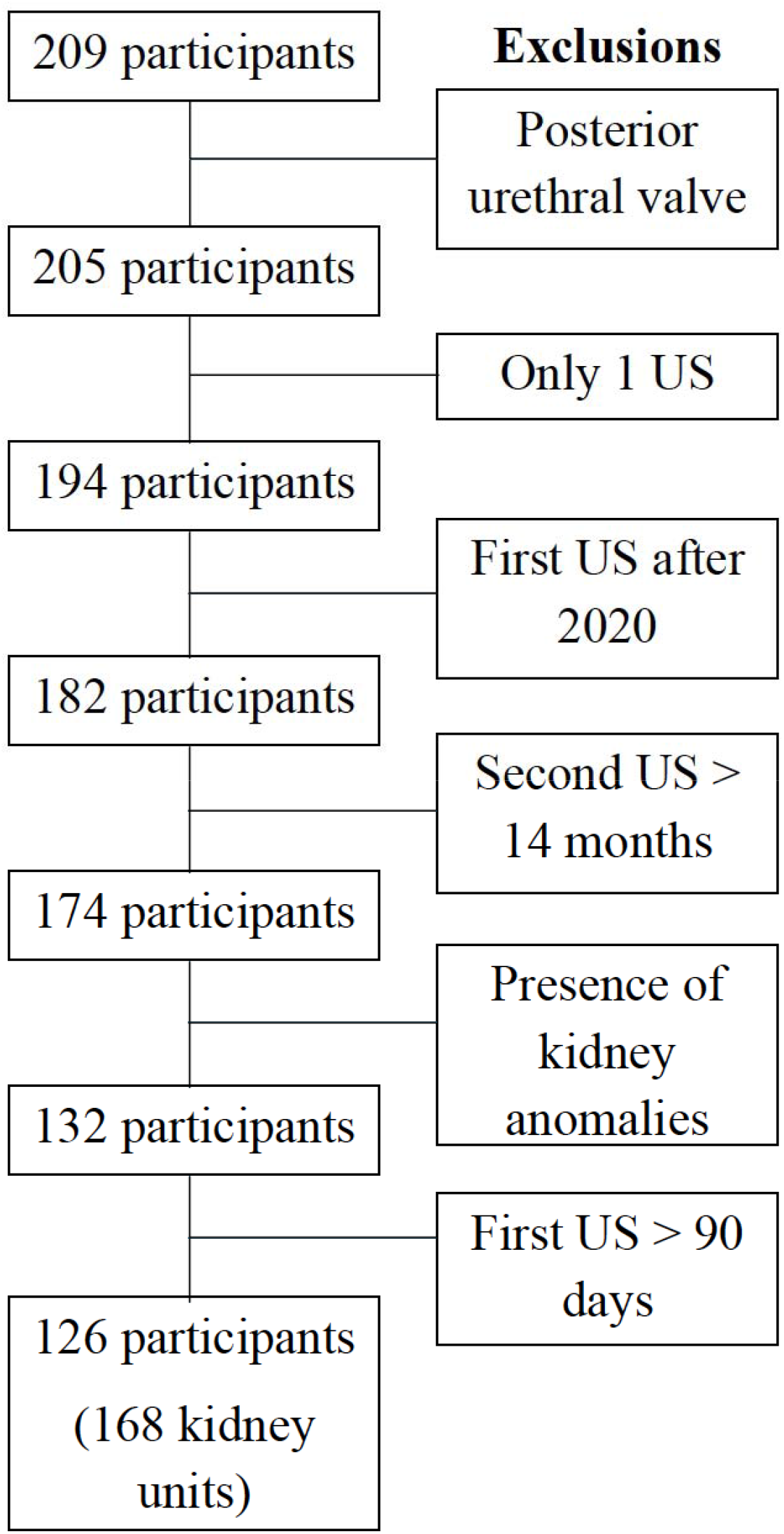
Patient cohort

**Table 1.** Baseline characteristics of the 126 children enrolled in the study.

| **Characteristic** | **N = 126<sup>†</sup>** |
| --- | --- |
| Age (Days), median (IQR) | 12 (8, 23) |
| Sex |  |
| Female | 28 (22%) |
| Male | 98 (78%) |
| Number of kidneys with hydronephrosis |  |
| 1 | 84 (67%) |
| 2 | 42 (33%) |

**Table 2.** Cox proportional hazards, N = 168, number of events = 81.

| **Characteristic** | **N** | **HR** | **95% CI** | **p-value** |
| --- | --- | --- | --- | --- |
| Grade rounded up |  |  |  | <0.001 |
| 1 | 23 | — | — |  |
| 2 | 70 | 0.59 | 0.34, 1.01 | 0.054 |
| 3 | 45 | 0.23 | 0.12, 0.47 | <0.001 |
| 4 | 30 | 0.04 | 0.01, 0.14 | <0.001 |
| Sex |  |  |  | 0.6 |
| Female | 37 | — | — |  |
| Male | 131 | 0.87 | 0.53, 1.45 | 0.6 |
| Age at first US | 168 | 1.01 | 0.99, 1.03 | 0.2 |

Abbreviations: CI = Confidence Interval, HR = Hazard Ratio

### Resolution of Congenital Hydronephrosis

The median duration of follow up (time to natural resolution or end of follow up) was 12.5 (IQR 5.9, 24.3) months. In terms of resolution at a kidney unit level, there were 81/168 (48%) kidney units that resolved spontaneously with equal distribution by side (Figure 2). A total of 49/105 (47%) kidney units resolved on the left side and 32/63 (51%) kidney units resolved on the right side. In an unadjusted analysis (Figure 3), low-grade hydronephrosis resolves sooner on average than high-grade hydronephrosis (P<0.0001). In the low-grade hydronephrosis group (SFU grade 1-2), the estimated proportion resolved at 24 months was 69% (95% Confidence Interval (CI) 54%, 78%), while in the high-grade hydronephrosis group (SFU grade 3-4), the estimated proportion resolved was 24% (95% CI 11%, 35%).

**Figure 2.**
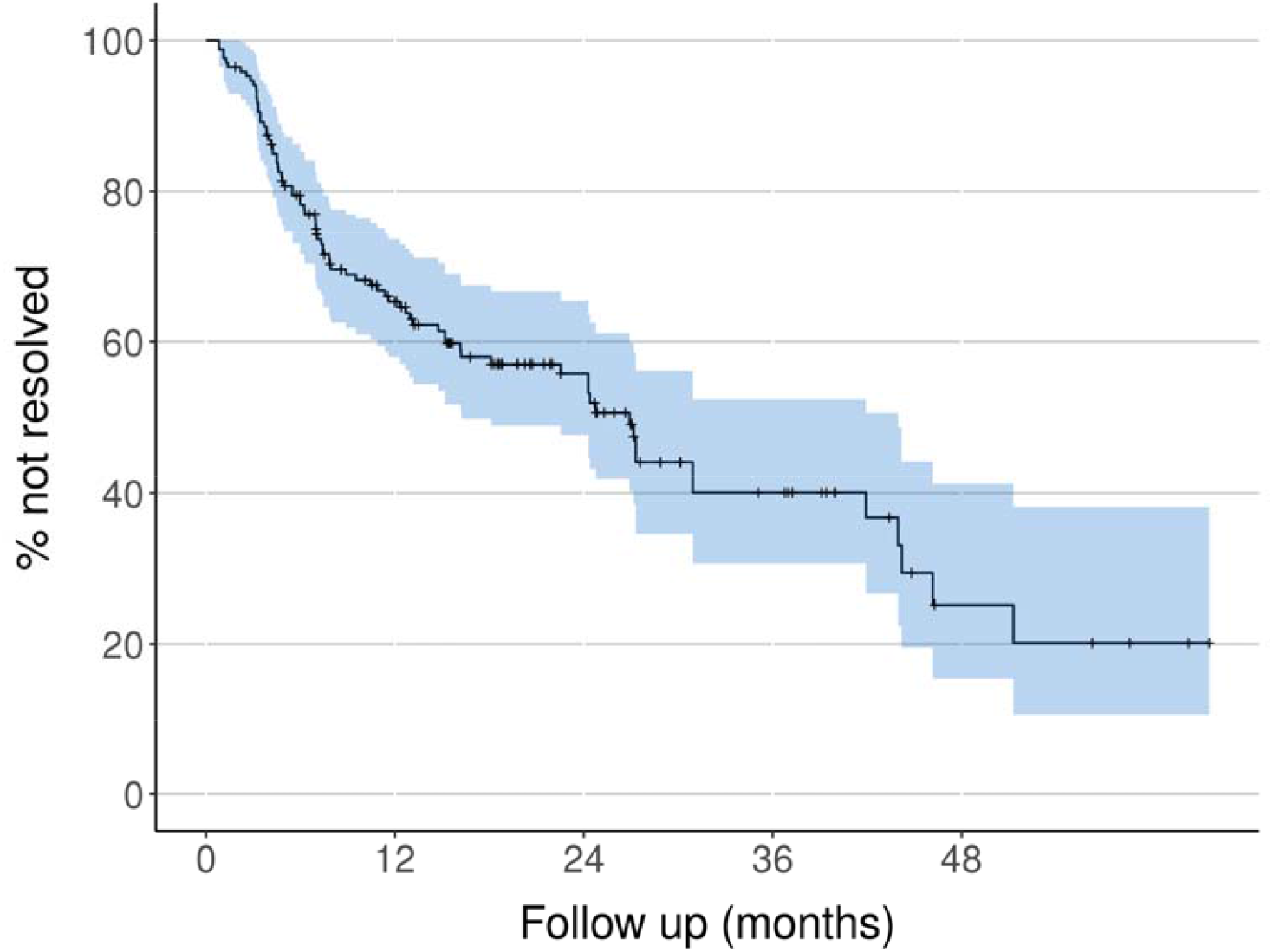
Kaplan-Meier curve showing time to resolution adjusted for clustering by participant (months).

**Figure 3.**
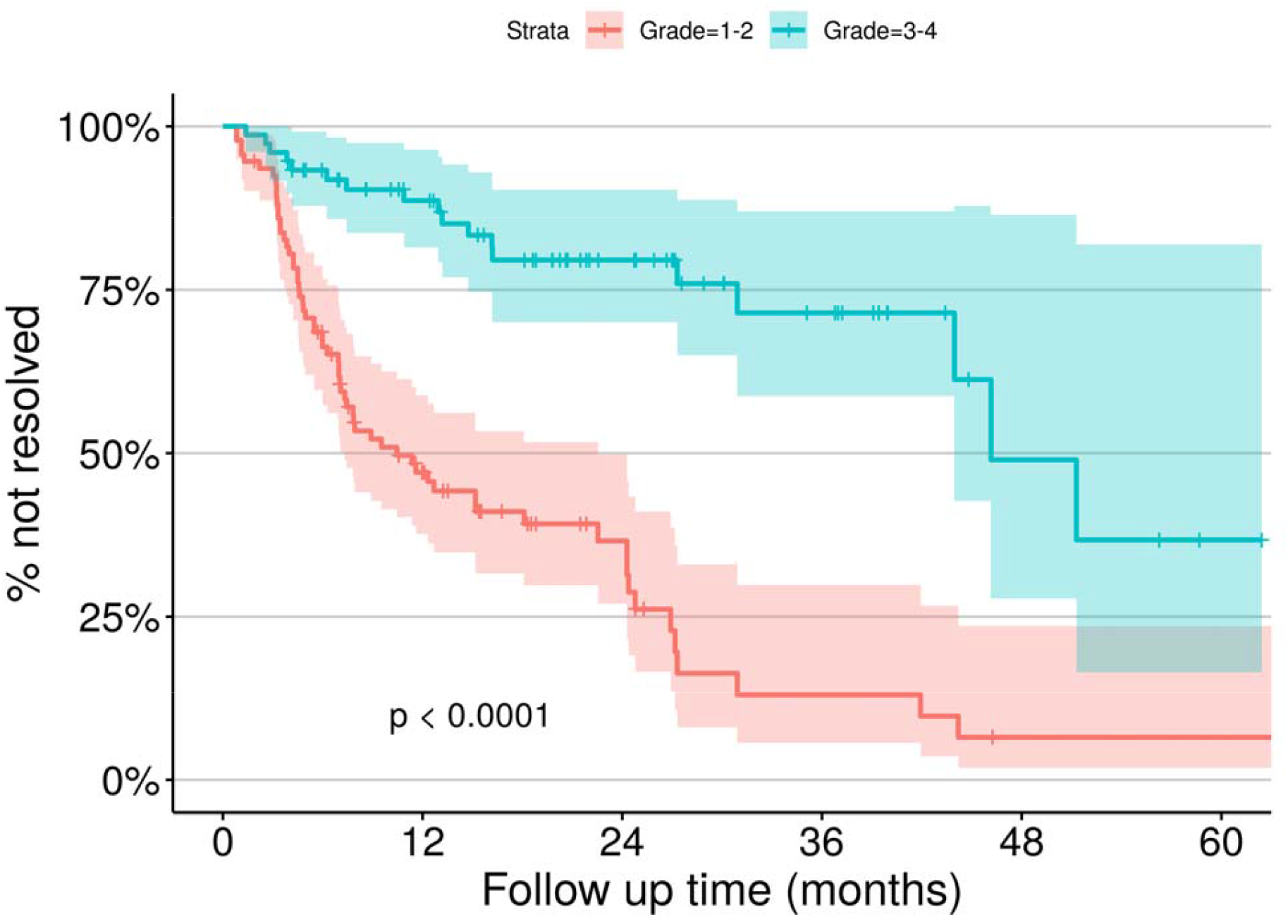
Kaplan-Meier curves by initial grade showing time to resolution adjusted for clustering by participant.

In multivariable Cox proportional hazards analysis, after adjusting for age at initial ultrasound, and sex, there was an association between initial SFU grade and time to natural resolution. Specifically, when compared to grade 1, adjusted HR for grades 2, 3 and 4 were 0.59 [95% CI 0.34-1.01], 0.23 [95% CI 0.12,0.47] and 0.04 [95% CI 0.01,0.14], respectively; p<0.001). These findings were unchanged in sensitivity analyses excluding kidney units that had undergone surgical intervention (Supplemental Table 1) and when SFU grades that were reported as a range were rounded down (Supplemental Table 2).

## Discussion

This retrospective study highlights that high-grade congenital hydronephrosis resolves more slowly than low-grade. This finding is consistent with a study conducted by Longpre et al., which aimed to determine independent predictors of resolution in patients with antenatal hydronephrosis [2]. In 118 kidney units (100 children), 62 resolved, 27 persisted and 29 underwent pyeloplasty. The authors identified that an initial larger anterior-posterior diameter (APD) was predictive of surgical intervention [2]. The study concluded that the APD on kidney ultrasound and SFU grade 4 independently predicted a lower likelihood of resolution of antenatal hydronephrosis [2]. Our study revealed similar findings in that SFU grade 4 was least associated with natural resolution. However, in our study, we observed that even those with SFU grade 3 were unlikely to resolve spontaneously. This observation allows for more personalized approaches to counseling families on the potential for resolution. In another study by VanDervoort et al., the authors aimed to determine the time course until spontaneous resolution of neonatal hydronephrosis and define risk factors associated with persisting lesions [4]. They reported a strong correlation between severity of hydronephrosis and likelihood of spontaneous resolution [4]. However, hydronephrosis was graded in terms of degree of severity (mild-moderate-severe) and did not include information related to SFU grading [4]. Of those with spontaneously resolving CH, approximately 10% had transient worsening at some time during their follow-up [4]. Interestingly, the authors identified that prematurity, urinary tract infections and perinatal complications did not affect the likelihood of resolution [4].

In contrast to the study from Longpre et al., where 24 children (25%) underwent open pyeloplasty for either progression of hydronephrosis, reduced and/or worsening differential function, or clinical complications such as pain and UTI, our study reports a minority of patients requiring surgical intervention (9 patients). A total of 5 patients underwent open pyeloplasty and 1 patient underwent a total nephrectomy. Reasons to proceed with surgical intervention included reduced differential kidney function or prolonged washout half-life time on renogram. This finding is in keeping with a study from O’Flynn et al. which demonstrated that patients with CH can be managed conservatively in the majority of cases, with 87% of their study population showing resolution [1]. Furthermore, this study demonstrated that no patient conservatively managed had any deterioration of their kidney function [1]. A subset of 7 patients had a differential kidney function of <40% on the affected side but subsequently their kidney function normalized [1].

Two studies were conducted in patients with only severe hydronephrosis: one with patients having severe unilateral hydronephrosis and the other study with patients who had severe bilateral hydronephrosis [6, 7]. Ulman et al. investigated the long-term follow-up of 104 patients with antenatally diagnosed severe unilateral hydronephrosis and concluded that the majority of patients with hydronephrosis, even severe (SFU grade 3 and 4) can be safely managed conservatively [6]. In patients who underwent pyeloplasty, the decision to intervene surgically was based on subsequent imaging rather than the initial severity of hydronephrosis or the differential kidney function [6]. Many patients who had reduced kidney function and did not undergo pyeloplasty eventually improved to have normal kidney function with resolving hydronephrosis [6]. The study specifically highlights the need for close follow-up during the first two years of life to identify children at highest risk of obstruction [6]. A later study by Onen et al. investigated patients with severe bilateral hydronephrosis [7]. Pyeloplasty was required in 13 kidneys (9 patients) out of 38 kidneys (19 patients) with evidence of obstructive injury, such as deterioration in their differential kidney function defined as a decline greater than 10% or progression of the hydronephrosis [7]. Current testing modalities cannot differentiate patients with high grade lesions that can be managed conservatively with close observation from those patients that will require pyeloplasty. Serial imaging with close follow-up is required to identify patients most at risk [7].

Our study is not without limitations. Given that our study is retrospective in nature, follow-up visits were not consistently at the same interval. Furthermore, while we had intended to look at other ultrasound markers, data retrieval issues precluded this. Ultrasound findings and SFU grades were reported by radiologists at our centre and could be predisposed to subjective interpretation. In some instances, hydronephrosis was reported to be between two SFU grades (example SFU grade 1-2). To mitigate the impact of misclassification of findings based on SFU grades, ultrasound findings between two grades were rounded to the highest SFU grade. We also conducted a sensitivity analysis using SFU grades that were rounded down to the lower SFU grade and obtained similar findings. Another limitation of our study is the inclusion of patients with vesicoureteral reflux. Given the invasive nature of a VCUG, current guidelines recommend performing a VCUG for patients with high-grade hydronephrosis or patients with low-grade hydronephrosis who develop a urinary tract infection [3]. Since not all patients in our cohort underwent a VCUG, patients with reflux could not all be reliably identified. In the time-to-event analyses, individuals who had a surgical intervention were censored at the time of surgery. This censoring may be information which could create a bias because the individuals for whom a surgical intervention was chosen are less likely to resolve naturally. However, this only applied to 12 kidney units and a sensitivity analysis excluding these units showed nearly identical results.

## Conclusion

The study attempted to identify factors associated with natural resolution of CH at a single centre. When considering all grades of CH together, resolution of CH is approximately 50%. Resolution of CH is similar between left and right kidneys. There is a significant difference in time to resolution between low grade (SFU 1, 2) and high grade hydronephrosis (SFU 3, 4). In comparison to Longpre et al., our study revealed that even SFU grade 3 CH resulted in prolonged time to natural recovery. In this study, a minority of patients required surgical interventions with most patients managed conservatively with close observation. Our data appears to support that the most critical period for monitoring is the first 2 years. Further research is necessary to identify biomarkers to assess obstruction and kidney damage that would allow prediction of need for surgical intervention in children with CH.

The authors declare no conflicts of interest.

Data is available upon reasonable request.

The authors received no funding to conduct this research.

## Data Availability

Data is available upon reasonable request.

## Supplemental Tables

**Supplemental Table 1.** Model with SFU grades rounded up and removal of surgical patients (sensitivity analysis) Cox model (N = 156, number of events = 81)

| Characteristic | N | HR | 95% CI | p-value |
| --- | --- | --- | --- | --- |
| Grade rounded up |  |  |  | <0.001 |
| 1 | 23 | — | — |  |
| 2 | 69 | 0.59 | 0.34, 1.02 | 0.057 |
| 3 | 44 | 0.24 | 0.12, 0.47 | <0.001 |
| 4 | 20 | 0.04 | 0.01, 0.16 | <0.001 |
| Sex |  |  |  | 0.6 |
| Female | 34 | — | — |  |
| Male | 122 | 0.87 | 0.53, 1.44 | 0.6 |
| Age at first US | 156 | 1.01 | 0.99, 1.03 | 0.2 |

Abbreviations: CI = Confidence Interval, HR = Hazard Ratio

**Supplemental Table 2.** Model with SFU grades rounded down (sensitivity analysis) Cox model (N = 168, number of events = 81)

| Characteristic | N | HR | 95% CI | p-value |
| --- | --- | --- | --- | --- |
| Grade rounded down |  |  |  | <0.001 |
| 1 | 44 | — | — |  |
| 2 | 73 | 0.58 | 0.37, 0.90 | 0.016 |
| 3 | 25 | 0.14 | 0.05, 0.41 | <0.001 |
| 4 | 26 | 0.05 | 0.01, 0.18 | <0.001 |
| Sex |  |  |  | 0.2 |
| Female | 37 | — | — |  |
| Male | 131 | 0.75 | 0.47, 1.19 | 0.2 |
| Age at first US | 168 | 1.02 | 0.99, 1.04 | 0.2 |

Abbreviations: CI = Confidence Interval, HR = Hazard Ratio

